# COVID19 epidemic growth rates have declined since early March in U.S. regions with active hospitalized case surveillance

**DOI:** 10.1101/2020.08.19.20178228

**Authors:** Rajiv Bhatia

**Affiliations:** Assistant Professor of Medicine (affiliated), Stanford University Physician, Palo Alto VA Health System 3801 Miranda Ave, Palo Alto CA 94304

## Abstract

**Introduction:** Optimal pandemic monitoring and management requires unbiased and regionally specific estimates of disease incidence and epidemic growth.

**Methods:** I estimated growth rates and doubling times across a 22-week period of the SARS-COV-2 pandemic using hospital admissions incidence data collected through the US CDC COVID-NET surveillance program which operates in 98 U.S. counties located in 13 states. I cross validated the growth measures using mortality incidence data for the same regions and time periods.

**Results:** Between March 1 and August 8, 2020, two distinct waves of epidemic activity occurred. During the first wave in the COVID-NET monitoring regions, the harmonic mean of the maximum weekly growth rate was 534% (Median: 575; Range: 250 to 2250) and this maximum occurred in the second or third week of March in different regions. The harmonic mean of the minimum doubling time occurred with maximum growth rate and was 0.35 weeks (Median 0.36 weeks; Range: 0.22 to 0.55 weeks). The harmonic mean of the maximum incidence rate during the first wave of the epidemic was 8.5 hospital admissions per 100,000 people per week (Median: 9.2, Range: 4 to 40.5) and the peak of epidemic infection transmission associated with this maximum occurred on or before March 27, 2020 in eight of the 13 regions. Dividing the 22-week observed period into four intervals, the harmonic mean of the weekly hospitalization incidence rate was highest during the second interval (4.6 hospitalizations per week per 100,000), then fell during the third and fourth intervals. Growth rates declined from 101 percent per week in the first interval to 2.5 percent per week in the last. Doubling time have lengthened from 3/5^th^ of a week in the first interval to 12.5 weeks in the last. Period by period, the cumulative incidence has grown primarily in a linear mode. The mean cumulative incidence of hospitalizations on Aug 8^th^, 2020 in the COVID-NET regions is 96 hospitalizations per 100,000. Regions which experienced the highest maximum weekly incidence rates or greatest cumulative incidence rates in the first wave, generally, but not uniformly, observed the lower incidence rates in the second wave. Growth measures calculated based on mortality incidence data corroborate these findings.

**Conclusions:** Declining epidemic growth rates of SARS-COV-2 infection appeared in early March in the first observations of nationwide hospital admissions surveillance program in multiple U.S. regions. A sizable fraction of the U.S. population may have been infected in a cryptic February epidemic acceleration phase. To more accurately monitor epidemic trends and inform pandemic mitigation planning going forward, the US CDC needs measures of epidemic disease incidence that better reflect clinical disease and account for large variations in case ascertainment strategies over time.

## Introduction

Reliable information on disease incidence is essential for epidemic monitoring and control. Disease incidence is the basis for guaging epidemic acceleration and decline and for informing decisions about the timing and intensity of disease control measures. Survey methods, laboratory methods, syndromic surveillance, hospitalizations, and deaths are all potentially useful and complementary sources of epidemic incidence data.

The type and quality of disease of incidence data reflects deliberative choices made by public health authorities as well as choices made by clinical professionals on whom disease surveillance programs depend and choices made by people who seek care.^1^ For example, choosing how to define a case and who to test influences observed case incidence.^2^ In a novel epidemic, each choice affecting incidence data can vary significantly over time. These choices are instrumental not only for disease control, but they also define the narrative of the epidemic.

The US CDC has been using cases, deaths and hospitalizations along with laboratory testing data to monitor the SARS-COV-2 pandemic. In the United States, case counts appear to be the primary public facing measure of epidemic incidence. During the first months of the epidemic, being a case required having a known exposure and presenting with compatible clinical disease. On April 5^th^, 2020 CSTE adopted a surveillance definition for coronavirus disease based on confirmatory laboratory evidence removing requirements for clinical illness.^3^ Epidemic incidence curves based on laboratory-confirmed cases thus no longer represent people who are sick and incidence has become sensitive to testing strategies and population test-seeking behaviors. At the same time, over the epidemic period, the US CDC’s testing strategy has evolved from restricting testing to narrow group of clinical and public health priorities to endorsing community testing of asymptomatic healthy individuals without medical encounters.^4^ This wide variation in the application of laboratory testing will limit both inter-regional and inter-epidemic time period comparisons.^5^

Measures of SARS-COV-2 fatalities are equally vulnerable to biases. WHO published an emergency use diagnostic code for SARS-COV-2 disease on April 24^th^, 2020 defining death due to COVID-19 as “a death resulting from a clinically compatible illness, in a probable or confirmed COVID-19 case, unless there is a clear alternative.”^6^ Explicit official diagnostic criteria associated with this ICD-10 code do not exist, The CDC’s advises medical practitioners to report COVID–19 on a death certificate based on their judgement if there is no confirmatory laboratory testing.^4^

Hospital admissions might provide a timely and less problematic data source for tracking pandemic disease incidence;^5^ however, U.S. surveillance definitions for hospitalized cases do not include clinical or diagnostic criteria. The widespread practice of screening asymptomatic hospital inpatients means that incidence data based on hospitalized cases is not free of bias.

US CDC investigators have been conducting active case surveillance for laboratory confirmed SARSCOV-2 associated hospital admissions through the COVID-NET program since March 1, 2020.^7^ ^8^ This monitoring network includes 98 counties in 13 participating U.S. states representing approximately about 9 percent of the U.S. population. The stated purpose of the COVID-NET program is “to provide weekly, population-based estimates of SARS-CoV-2-associated hospitalizations to inform the public health response.” The CDC makes age-stratified weekly hospitalization rates publicly available; however, as of this date, CDC investigators have not published measures of epidemic dynamics based on this hospitalization data.

Here, I compute and analyze cumulative incidence growth rates and doubling times using COVID-NET hospitalization monitoring data from March 1 through August 8, 2020. I corroborate the measures using mortality incidence data for the same regions and time periods.

## Data and Methods

I accessed weekly age-stratified hospitalization incidence rates from the U.S. CDC COVID-NET program URL.^9^ The 98 current COVID-NET counties are located in California, Colorado, Connecticut, Georgia, Maryland, Michigan, Minnesota, New Mexico, New York, Ohio, Oregon, Tennessee, and Utah. These regions have been active participants in either the Emerging Infections Program (EIP) and the Influenza Hospitalization Surveillance Project (IHSP). Individual participating counties are listed in the supplemental materials.

I provide the detailed COVID-NET study protocol as supplementary materials. Briefly, investigators defined a case as a hospitalized resident of the participating counties who had evidence of a positive SARS-CoV-2 test less than 14 days prior to admission and less than three days after admission. Hospitalized patients who had a positive test more than 3 days after admission and did not have respiratory symptoms during this time period were considered hospital acquired cases. The study protocol did not apply an additional clinical criterion (e.g., pneumonia, fever, systemic inflammatory response) or a criterion for a COVID-19 specific ICD-10 code to establish case status.

COVID-NET hospitalization incidence rates represent the number of residents of a defined area who are hospitalized with a positive SARS-CoV-2 laboratory test divided by the total population within that defined area. Public data include disaggregated incidence and cumulative incidence rates for the 0–17, 18–49, 50–64, 65–74, 75–84, 85+ years old age groups.

After accessing the data and plotting the weekly incidence rates, I re-computed the cumulative incidence rate (CR) as the cumulative sum of weekly incidence rates. I then computed the growth rate as the week-to-week change in cumulative incidence (GR = CR/ lag (CR) –1). I estimated the doubling time of cumulative incidence at each weekly observation assuming a constant growth rate within each week period (DT = ln (2) / ln (1 + GR).

In order to represent the timing of these observations as those for infection incidence, I adjusted the date of the observations for the mean time interval from infection to observed hospital admission. Specifically, I combined the 6 days estimated mean interval from infection to symptoms and the 6 day median interval between symptoms and hospital admission.^10^ To assess early epidemic growth, I enumerated the day at which maximum incidence and maximum growth rates occurred during the first half of observations. In order to assess trends over the observed period, I divided the data into four equal time periods and computed the harmonic means of each interval’s measures. I provide the computed data by period in tables in the supplemental materials.

To corroborate these hospital admission-based growth measures, I replicated methods using publicly available county-level COVID-19 death incidence.^11^ I summed the incident deaths attributed to counties included in each COVID-NET state region and calculated weekly SARS-COV-2 associated mortality incidence using U.S. census county population estimates for 2018. I then computed cumulative incidence, growth rates, and doubling times as above. To place the deaths at their approximate time of infection, I adjusted the observations by 28 days based on the aforementioned incubation period combined with interval estimates published by the CDC derived from the COVID-NET data set. (symptoms to death: median 15 days; death to reporting: median 7 days).

## Findings

Figure 1 (Panels A-D) illustrates the hospital admissions incidence rate, the cumulative incidence rate, the week-to-week epidemic growth rate, and the doubling time in weeks across the 22-week observation period in the COVID-NET monitoring regions. Figure S1, included in the supplemental materials, provides region-by-region plots of each incidence and growth measure across the observation period. Figures and plots reflect observations based on their estimated time of infection. Thus, I plot admissions observed during the 7-day period of Mar 1 to Mar 7 as infections occurring during the 7-day period of Feb 18 to 24.

Figure 1 Panel A illustrates the large variability in the magnitude of the hospitalization incidence rates among regions. There are two distinct waves of epidemic growth and decay – a first wave of beginning at the onset of observations and decaying by May and second, typically smaller wave observed by some regions in late June and July.

The harmonic mean of the regional maximum incidence rates during the period of the first wave is 8.5 hospital admissions per 100,00) per week (Median: 9.2, Range: 4 – 40.5). The first regional maximum of the associated infection incidence rate occurred by March 20^th^ in six regions and by March 27^th^, the median day, in eight of the thirteen regions. (Figure 2) Some states, namely Ohio, New Mexico, Minnesota, and Utah, experience their first peak of activity over a month later than states with early peaks, demonstrating decelerating growth up to these peaks.

**Figure 1.**
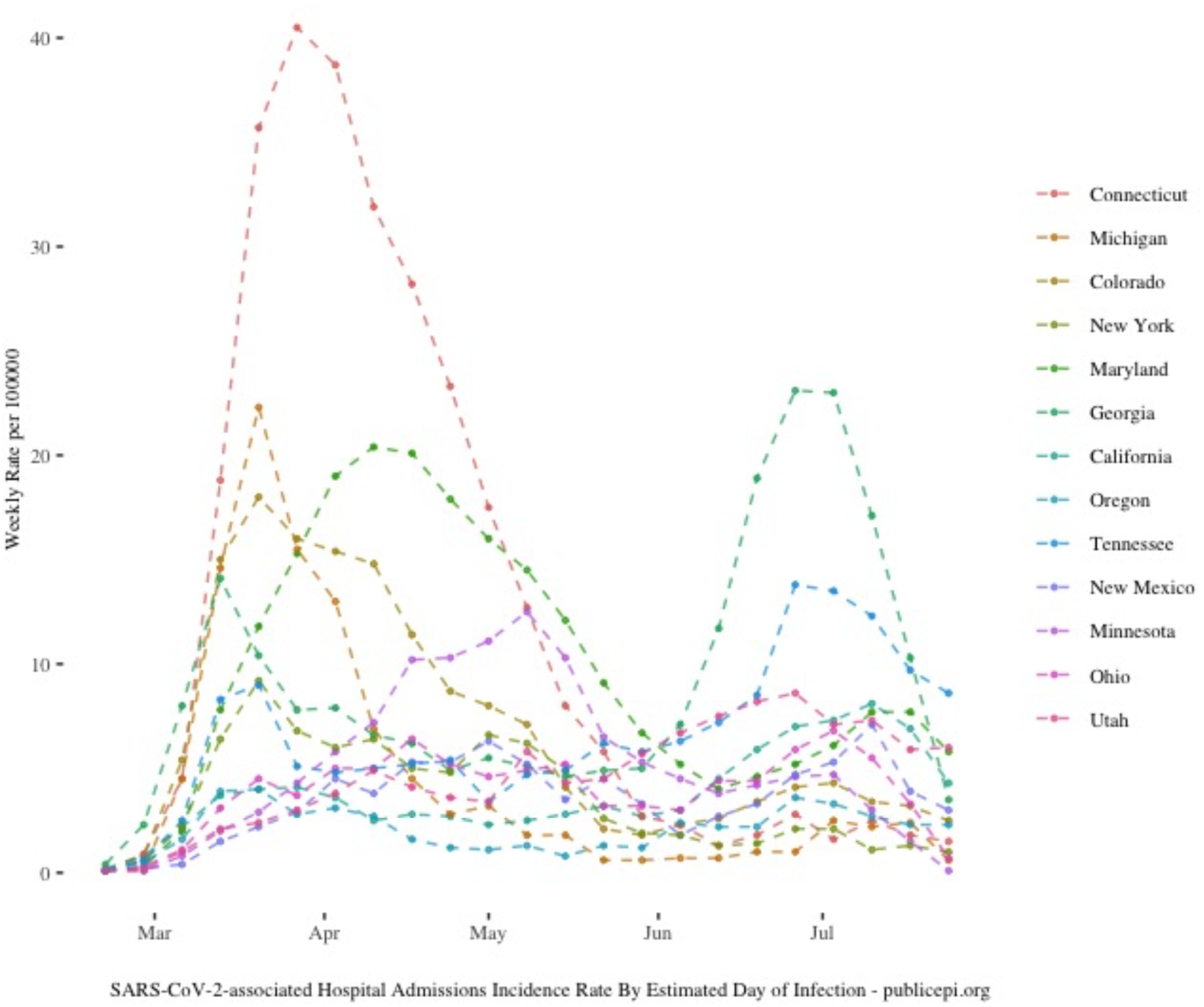
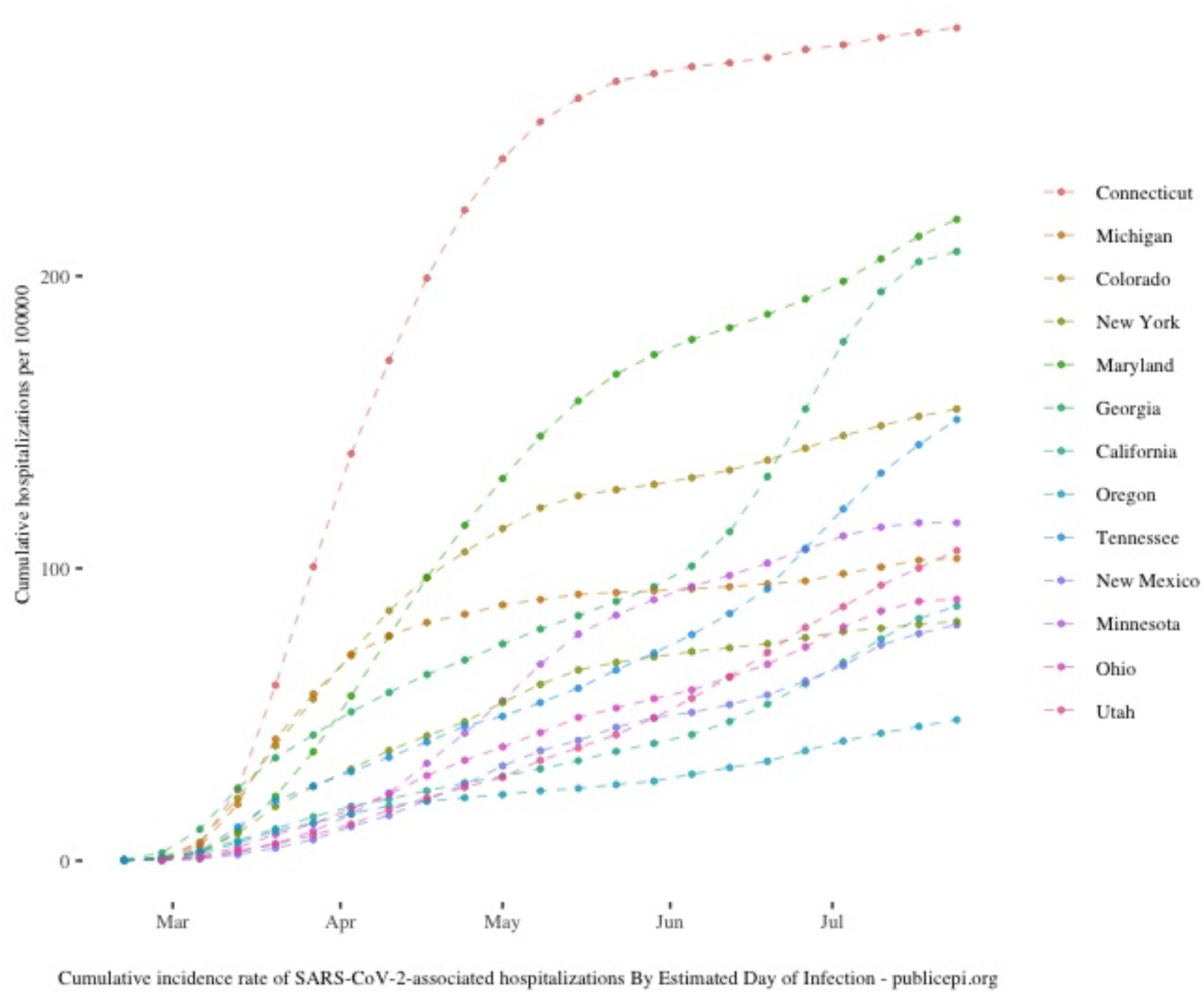
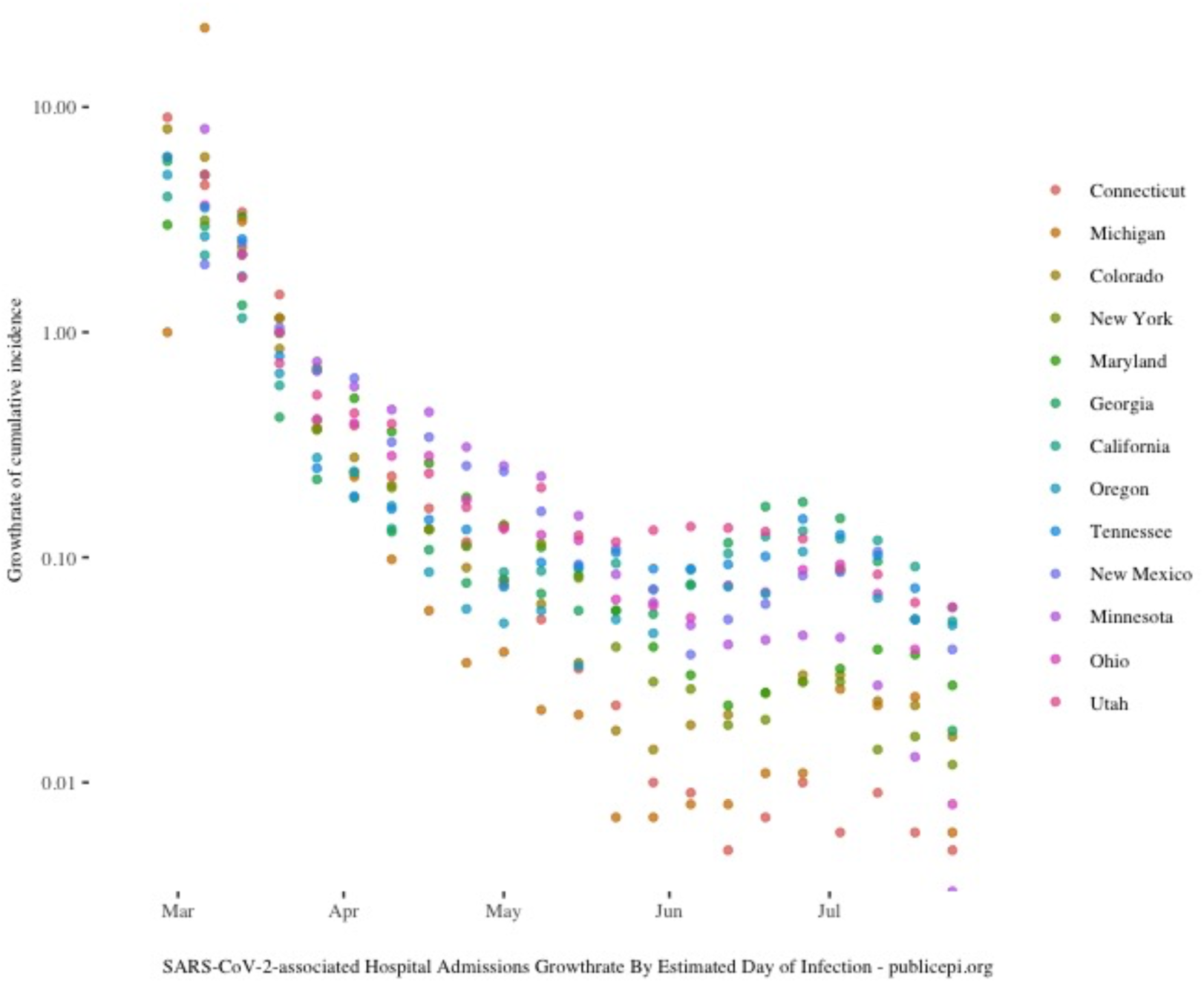
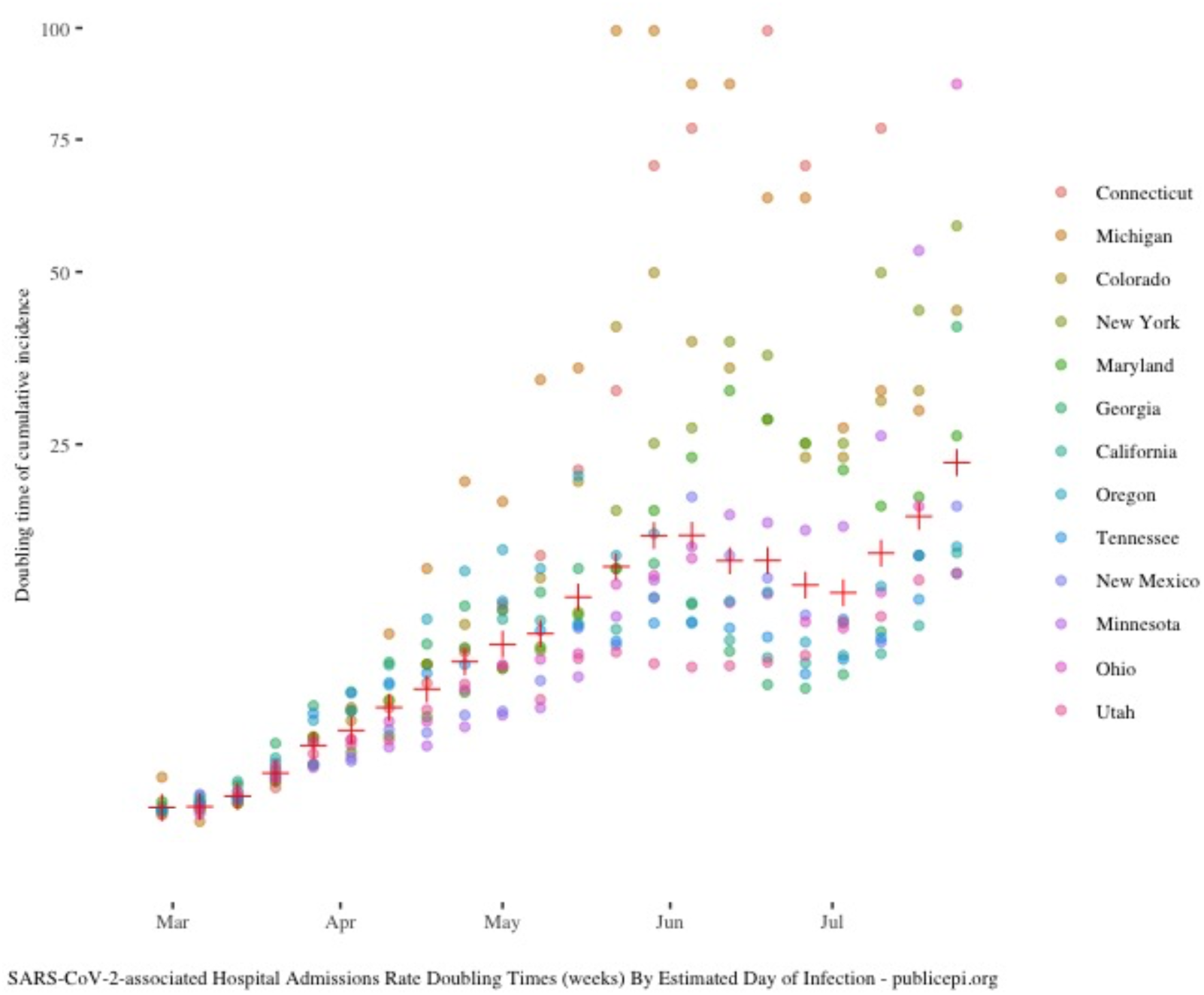
(Panel A-D) Epidemic period measures in 13 United States COVID-NET monitoring regions. Panel A. Weekly hospital admissions incidence; Panel B. Cumulative SARS-COV-2 associated hospitalization incidence in the 13 COVID-NET monitoring regions; Panel C. Week to week change in cumulative incidence (growth rates); Panel D. Doubling times in weeks. The red crosses are median values for each weekly observation.

**Table 1:** Maximum weekly hospital admission incidence rates per 100,000, cumulative incidence rate, and maximum cumulative incidence growth rate in COVID-NET regions observed between March 1 and May 23, 2020.

| Region | Maximum weekly hospital admissions Incidence | Est. day of maximum infection incidence rate | Period cumulative Incidence Rate | Maximum growth rate (% change) | Est. day of maximum infection growth rate |
| --- | --- | --- | --- | --- | --- |
| Connecticut | 40.5 | 3/27/20 | 252.8 | 900 | 2/28/20 |
| Michigan | 22.3 | 3/20/20 | 89.3 | 2250 | 3/6/20 |
| Colorado | 18 | 3/20/20 | 120.7 | 800 | 2/28/20 |
| New York | 9.2 | 3/20/20 | 60.3 | 600 | 2/28/20 |
| Maryland | 20.4 | 4/10/20 | 145.2 | 500 | 3/6/20 |
| Georgia | 14.1 | 3/13/20 | 79.2 | 575 | 2/28/20 |
| California | 4.1 | 3/27/20 | 31.4 | 400 | 2/28/20 |
| Oregon | 4 | 3/20/20 | 23.9 | 500 | 2/28/20 |
| Tennessee | 9 | 3/20/20 | 54.1 | 600 | 2/28/20 |
| New Mexico | 6.3 | 5/1/20 | 37.6 | 250 | 3/13/20 |
| Minnesota | 12.5 | 5/8/20 | 67.2 | 800 | 3/6/20 |
| Ohio | 6.4 | 4/17/20 | 43.8 | 366.7 | 3/6/20 |
| Utah | 5.8 | 5/8/20 | 34.3 | 500 | 3/6/20 |

In most regions, the plot of cumulative incidence rates (Figure 1b) shows an initial period of decelerating exponential growth followed by a period of linear growth. The harmonic mean cumulative incidence rate after the first wave in these regions is 53 hospitalizations per 100,000. (Median: 60.3; Range: 24–253) The calculated maximum growth rates are highest in Colorado, Connecticut, Michigan, and Minnesota. However, growth rates moderate faster in Colorado and Michigan than they do in Connecticut. Maryland, which begins at a lower growth rate than Colorado and Michigan, reaches a higher cumulative incidence than these two states by the midpoint of the observation period.

In the second half of this observation period, several regions with high incidence rates or high cumulative incidence in the first wave maintain predominantly linear growth while several regions with less intense first wave activity have relatively higher incidence rates. Several of these latter regions have a second period of epidemic acceleration.

**Figure 2.**
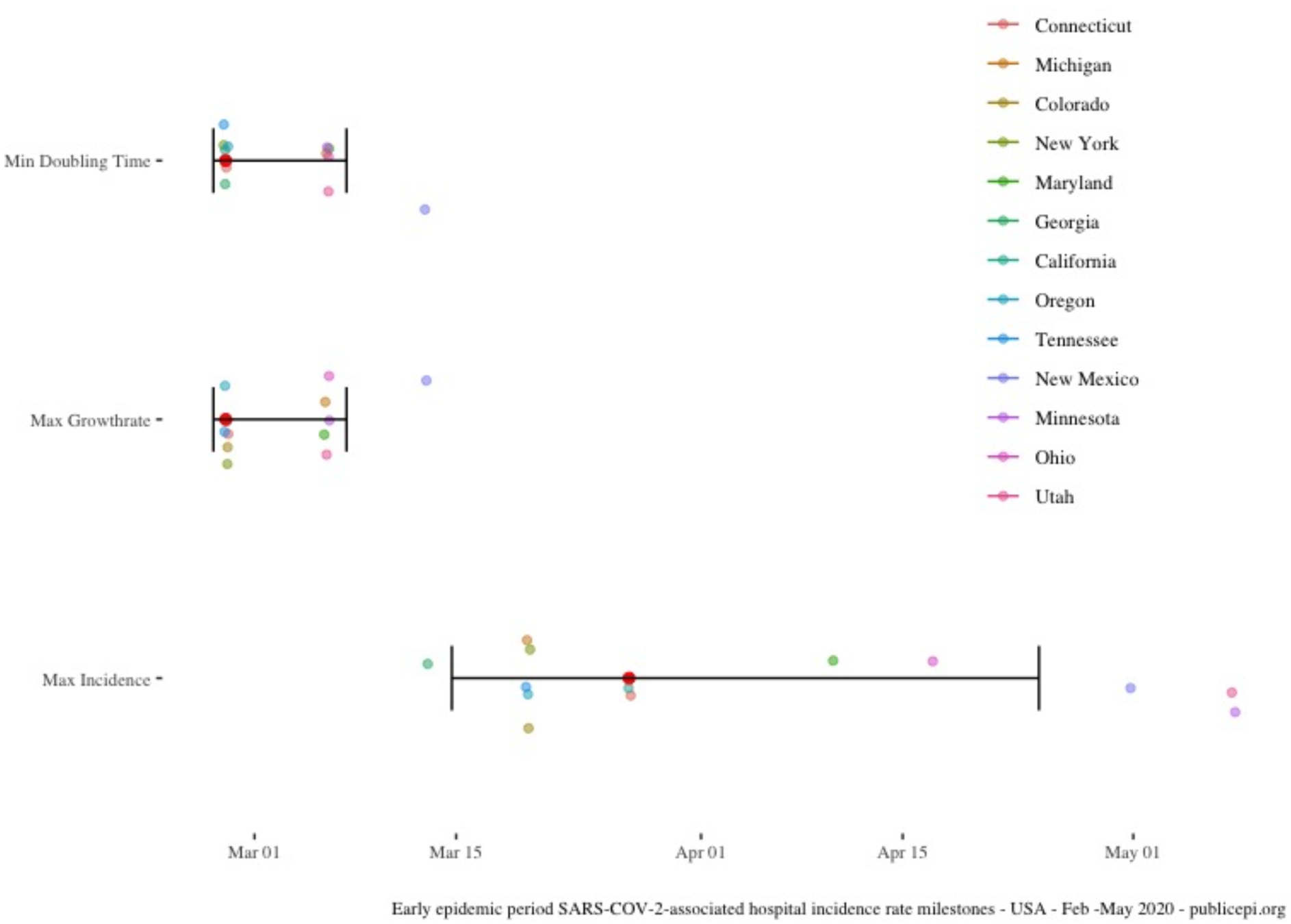
Timing of epidemic milestones in the first 10 weeks of hospital admissions monitoring. Both the growth rate maximum and the doubling time minimum fall at the beginning of the monitoring period. The median of the day of the maximum incidence falls on March 27^th^ with a cluster of regions experiencing maxima in the middle of March and another cluster in mid-April through May.

The plot of growth rates illustrates how the deceleration of the cumulative incidence curve begins in almost every region with the onset of observations. In the first wave, the harmonic mean of the maximum weekly regional growth rate is 534% (Median: 575; Range: 250 – 2250) and occurred during the first weeks of observations in most regions.

Growth rates decline from the initial observation in all regions from this initial maximum. Michigan and Maryland have a single week to week increase in the estimated growth rate from the second to third week of observation and then decline. New Mexico observes growth from the third to fourth week of observation. After April, growth rates diverge among regions with Connecticut, Michigan, New York, and Colorado on lower growth rate trajectories in the second half of the observed period.

The harmonic mean of the minimum doubling time, which occurs with the maximum growth rate, was 0.35 weeks (Median 0.36 weeks; Range: 0.22 – 0.55 weeks) The doubling time of the cumulative incidence progressively lengthens over the course of the epidemic period in all regions; however, the plot of doubling times also illustrates larger differences in epidemic growth among regions during the second half of the observation period. Several of the regions with the highest first period cumulative incidence including Connecticut, Colorado, Michigan, and New York have the longest doubling times at the end of the observation period. Some regions experience a period of shortening of doubling times after the beginning of June co-incident with second wave activity with a return to the trend of lengthening in July.

**Table 2.** Harmonic means of SARS-COV-2-associated hospitalization incidence rates per 100.000 people, cumulative incidence rates, growth rates and doubling times in COVID-NET monitoring regions between March and July 2020.

| Quarter | Period of Observed SARS-COV-2 Hospitalizations | Period of SARS-COV-2 Infections (estimated) | Weekly hospitalization incidence Rate | Cumulative Incidence Rate | Week to week growth rate (Pct. Change) | Doubling time (Weeks) |
| --- | --- | --- | --- | --- | --- | --- |
| First | Mar 1 – Apr 11 | Feb 18 – Mar 30 | 0.4 | 0.6 | 100.8 | 0.6 |
| Second | Apr 12 – May 23 | Mar 31 – May 11 | 4.6 | 36.4 | 12.0 | 4.1 |
| Third | May 24- Jul 4 | May 12 – Jun 22 | 2.6 | 67.1 | 2.9 | 11.6 |
| Fourth | Jul 5 – Aug 8 | Jun 23 – Jul 27 | 3.0 | 96.2 | 2.5 | 12.5 |

Table 2 enumerates the harmonic mean incidence rate, cumulative incidence, growth rate, and doubling time computed for four-time intervals in the observed epidemic period. For the 13 US monitoring regions, the harmonic mean of the weekly hospitalization incidence rate was highest during the second interval (4.6 hospitalizations per week per 100,000), fell and then remained steady during the third and fourth intervals. Growth rates declined from 101 percent per week in the first interval to 2.5 percent per week in the last. Doubling times lengthened from 3/5^th^ of a week in the first interval to 12.5 weeks in the last. Period by period, the cumulative incidence has grown in an approximately linear mode. The mean cumulative incidence of hospitalizations on Aug 8^th^, 2020 in the COVID-NET monitoring regions is 96 hospitalizations per 100,000.

Figure 3 illustrates the harmonic mean of the regional period-specific harmonic mean doubling times, demonstrating the consistent increase across periods and a progressively wider spread among regions.

**Figure 3.**
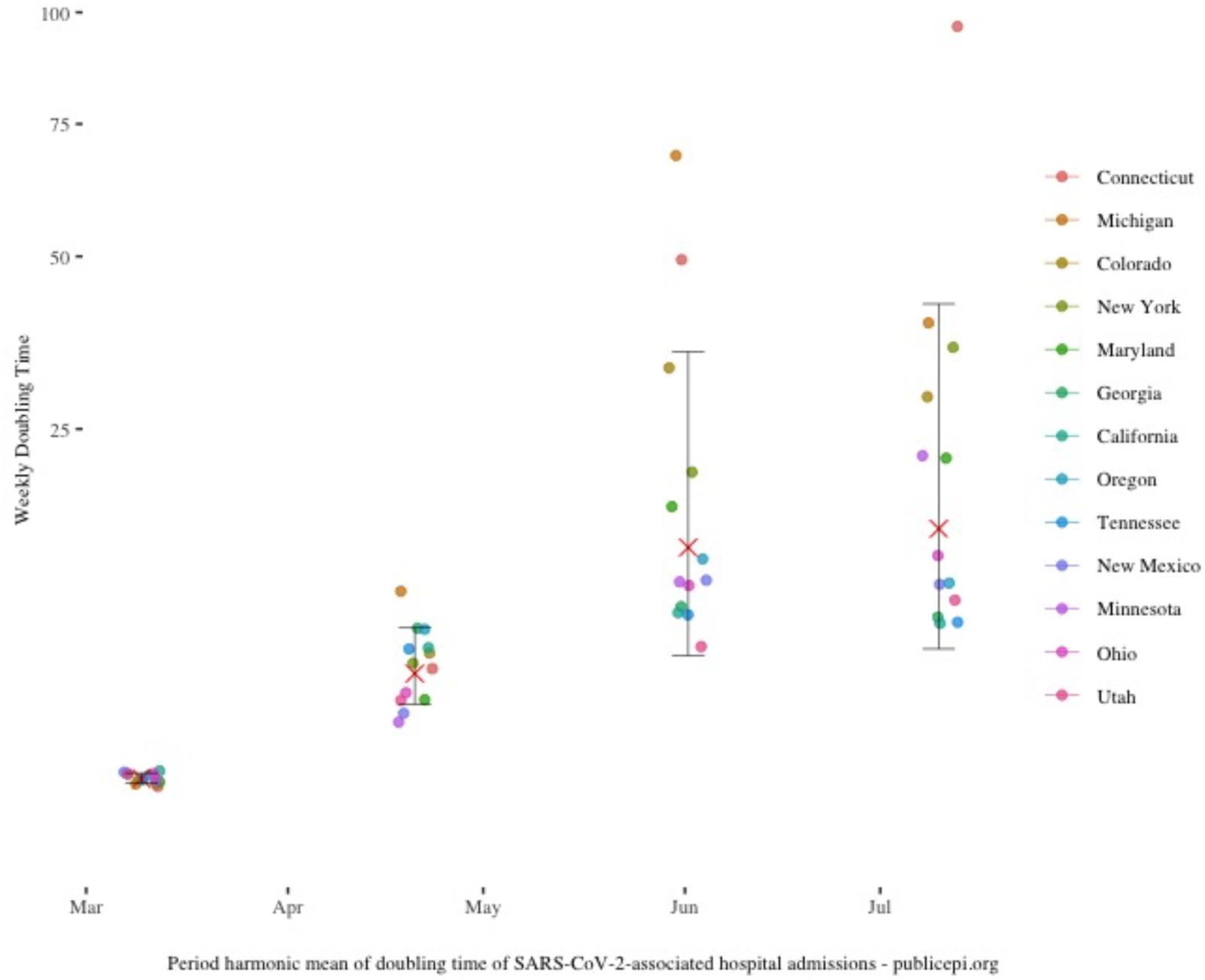
Harmonic means of regional doubling times with each of four equally spaced intervals within the current epidemic period. The error bar contains the 95% confidence interval of these values. The red X represents the harmonic mean of the period state means.

Figure 4 shows the cumulative SARS-COV-2 associated mortality incidence rate (Panel A) and the weekly cumulative mortality growth rates in the COVID-NET regions. (Panel B) Table 3 enumerates the harmonic mean mortality incidence rate, cumulative incidence, growth rate, and doubling time for all regions computed for the same four-time intervals as was done for the hospitalization data. Across four intervals of the observed epidemic period, measures of mortality incidence and cumulative mortality incidence growth demonstrate similar patterns to those estimated based on hospitalization incidence data. Across the 13 regions, mortality growth rates declined from the first interval and mortality incidence rates peaked in the second interval of observation. Based on mortality incidence data, SARS-COV-2 epidemic growth rates have declined in the COVID-NET regions since the first interval of observation.

**Table 3.**
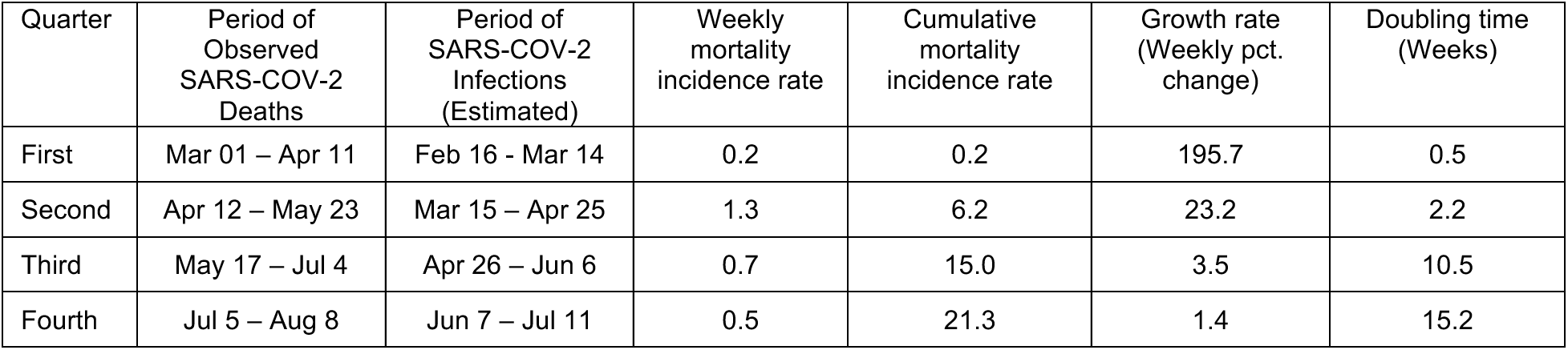
Harmonic means of SARS-COV-2-associated mortality incidence rates per 100,000 people, cumulative incidence rates, growth rates and doubling times in 13 COVID-NET monitoring regions between March and July 2020.

**Figure 4.**
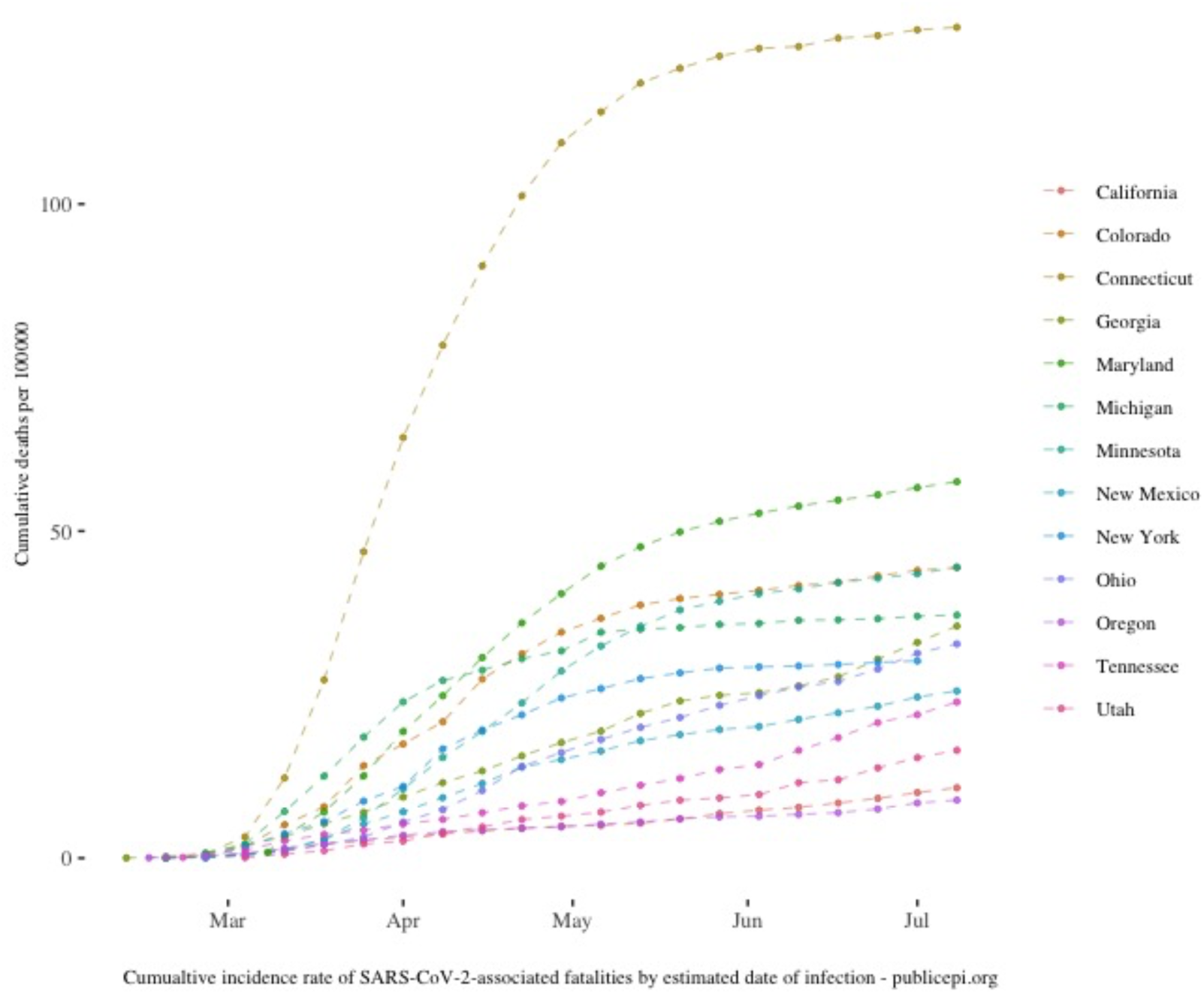
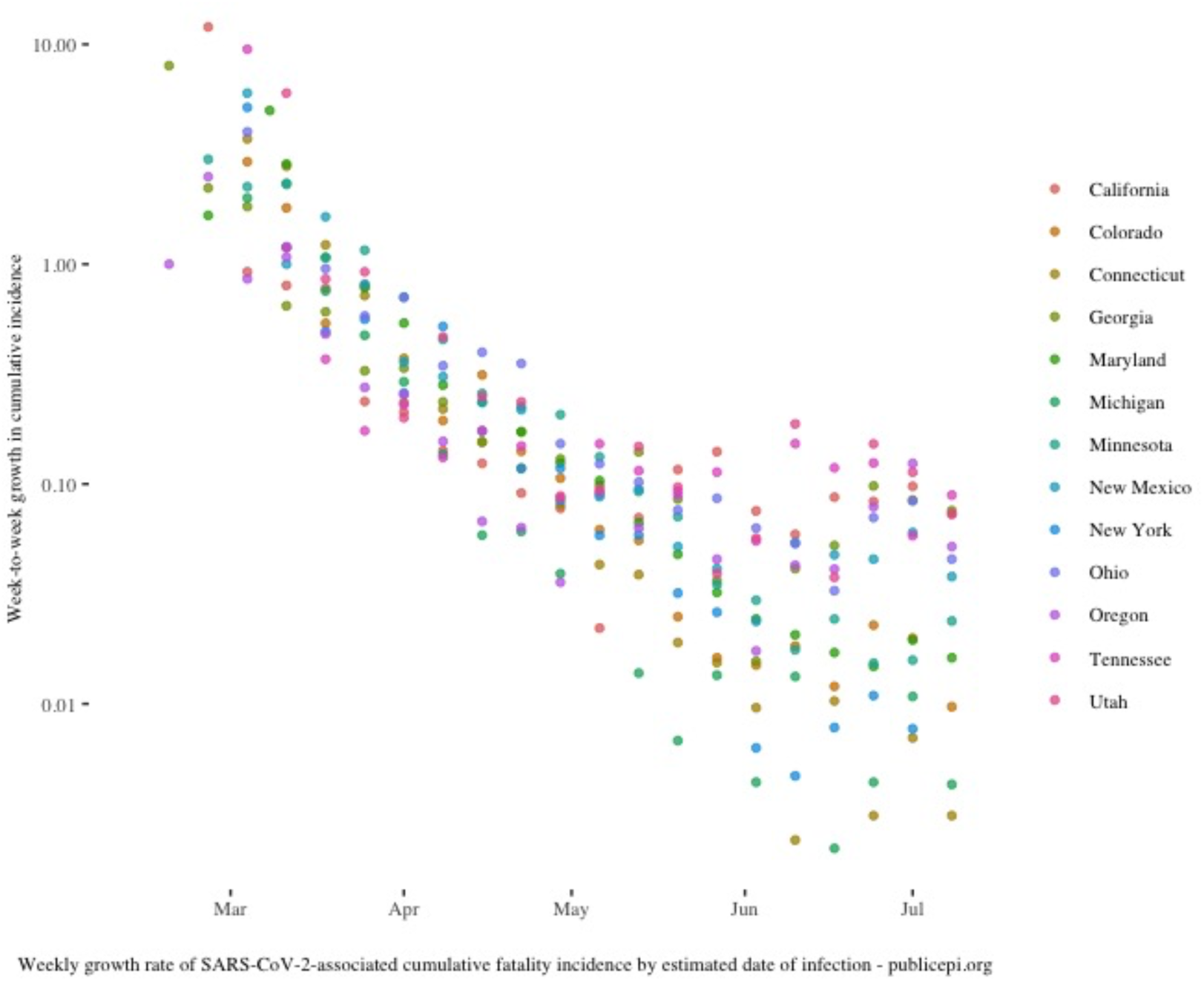
(Panel A-B) Epidemic period SARS-COV-2 fatality incidence measures in 13 United States COVID-NET monitoring regions. Panel A. Cumulative SARS-COV-2 associated mortality incidence. Panel B. Week to week growth rate in cumulative mortality incidence.

## Discussion

Data from both hospitalization and mortality incidence observations in U.S. CDC monitoring regions demonstrates that the SARS-COV-2 epidemic has been in a deceleration phase in many U.S. regions for most of the period of laboratory-based case ascertainment. The observation of maximum growth rates from first observations of hospital admissions along with the observations of maximum hospitalization incidence within weeks of the first observations support this conclusion. Analysis based on mortality incidence provides corroborating evidence.

The inability to observe accelerating growth in March 2020 suggests that, accounting for the infection to hospital admissions interval, an epidemic acceleration phase began before Feb 18, 2020. Others have come to similar conclusions. Analysis based on airline travel patterns suggests that early seeding through global travel and widespread domestic propogation led to epidemic initiation in the middle of February.^12^ Analysis of genomic data similarly concludes that SARS-COV- transmission networks became established in early to mid-February 2020.^13^ Furthermore, Pan American Health Organization Flu Net laboratory monitoring data shows increasing SARS-COV-2 test positivity early in the month of February in many countries in the Western Hemisphere.^14^ In Ecuador, for example, laboratory test positivity for SARS-CoV-2 increased from 1 to 30% from the beginning to the end of the month of February.

Evidence of pre-symptomatic and asymptomatic transmission along with a low clinical fraction further supports the plausibility of undetected epidemic acceleration. In January, Singapore health officials found asymptomatic individuals with RT-PCR tests positive for SAR-COV-2 among travelers returning from Wuhan,^15,16^ and identified evidence for pre-symptomatic transmission through contact tracing.^17^ Japanese researchers found asymptomatic infections among Japanese citizens evacuated from Wuhan, China^18^ and among those infected on the Diamond princess cruise ship.^19^ One Chinese modeling effort concluded that transmission from undocumented cases was responsible for 80% of the documented cases in Wuhan before their travel restrictions.^20^

In January and February 2020, the US CDC’s COVID-19 response team conducted surveillance activities including case-contact monitoring but did not test asymptomatic contacts or consider asymptomatic carriage among inbound travelers. In contrast, Singapore had enhanced surveillance at the end of January, testing all hospitalized patients with pneumonia as well as ambulatory patients with influenza-like illness.^21^ Though Singapore had few confirmed cases during this time period, time from symptoms to isolation for both international and domestic cases fell significantly after implementing this enhanced surveillance. Notably, the last assessment of this period by the US CDC response team maintains that only sporadic infection occurred in January and February –- the time when domestic RT-PCR testing was not available.^22^

If widespread epidemic community transmission did occur in February in the United States, children and young adults may have been the first to experience it. Based on analysis of the frequency and characteristics of contacts across age groups, Mossong argued that 5- to 19-year-olds are like to be the ones with the highest incidence of infection during the initial epidemic phase of an emerging infection transmitted person to person.^23^ Others have confirmed that children and young adults have higher baseline contact rates than adults and generally assort with people of their own age groups.^24^ For SARSCOV-2 specifically, the fraction of the young who appear to develop symptoms after laboratory-confirmed infection appears very low.^25^ For younger populations with symptoms, the non-specific clinical presentation may have allowed notable fraction of this populations to experience SARS-COV-2 infection without coming to medical attention. Even in hospitalized patients with pneumonia, the lack of identification of a specific respiratory pathogen is not unusual.^26^

Supporting February epidemic acceleration, in New York City, syndromic surveillance data shows an epidemic wave of influenza like illness (ILI) in the 5–17-year-old population in the last weeks of February not explained by other viral markers. The age-specific wave peaks about three weeks ahead of the peak in the city’s wave of SARS-COV-2 associated hospitalizations.^27^ Additionally, national data from sentinel Flu Net providers documents an unexplained increase of 32,000 total patient visits during the week of February 23–29, 2020 which occurred when influenza test positivity rates had been declining.^28^ Visits for influenza like illness, specifically, increased significantly during the following week. Taken together, the above observations along with those reported here, make widespread transmission of SARS-COV-2 during a cryptic February epidemic acceleration phase likely. (Table 4)

**Table 4.** Plausible explanations of declining epidemic growth rates in the early period or observed SARS-COV-2 hospital admissions surveillance

| **Explanation** | **Supporting observations** |
| --- | --- |
| Unconstrained cryptic epidemic initiation and acceleration in February | ▪ Pre-symptomatic and asymptomatic transmission.<br>▪ Symptom based disease surveillance.<br>▪ Low clinical fraction among children and young adults.<br>▪ February period growth of SARS-COV-2 laboratory test positivity in Western Hemisphere countries.<br>▪ Surge of sentinel provider and syndromic surveillance illness visits in late-February. |
| Under ascertainment of hospitalized cases | ▪ Non-specific illness presentations.<br>▪ Restrictive case identification criteria.<br>▪ Lack of diagnostic testing capacity.<br>▪ Lack of an operating syndromic case definition. |
| Societal level behavior change | ▪ Widespread increased social vigilance and hygiene (e.g., handwashing, hygiene, self-isolation)<br>▪ Voluntary social isolation (e.g., reductions in socializing, shopping and dining, travel, discretionary social engagements)<br>▪ Business risk management decisions (e.g. event cancellations, telework arrangements, conference cancellations, resort closures) |

The COVID-NET hospital admissions incidence data used in this study is has several limitations. Even though COVID-NET investigators used standard ascertainment methodology, the laboratory-based case definition did not require a compatible clinical syndrome nor physician’s diagnosis of COVID-19. Furthermore, testing for ascertain laboratory-positive SARS-COV-2 individuals has increased significantly across the observed epidemic period.

The various ascertainment biases that affect COVID-NET data are unlikely to change the conclusions here as they would have likely progressively increased ascertainment and thus observed growth rates during the month of March. Before February 28^th^, the CDC’s SARS-CoV-2 persons-under-investigation criteria required both compatible symptoms and close contact with a known case or travel within a high-risk country.^29^ On February 28th, the CDC broadened the criteria to include severe febrile pulmonary disease without an alternative explanation and without a known exposure to SARS-COV-2.^30^ On March 8^th^, CDC expanded SARS-COV-2 testing priorities to include hospitalized patients and symptomatic high-risk individuals.^31^ The latest CDC guidance advises the testing for asymptomatic individuals who subjectively suspect exposure.^4^ More recently, many jurisdictions in the US, have promoted “testing-ondemand” for asymptomatic individuals for any reason without a medical evaluation.^32^

Changes in testing rates have a large impact on incidence rates.^33^ In the first part of March, FDA had only authorized public health and clinical labs to conduct SARS-COV-2 tests. Figure 4 illustrates the weekly SARS-COV-2 testing rates in the US from March through July based on data reported by labs to the US CDC. In the first weeks of March, during the time observed hospital incidence rates grew, testing frequency increased six-fold, yet test positivity did not change. Increasing ascertainment may have accounted for some of the observed growth in hospitalized cases. More routine RT-PCR testing for hospitalized patients without SARS-COV-2 clinical syndromes may have contributed to the growth in incidence of hospitalized cases over the observed period.

**Figure 5.**
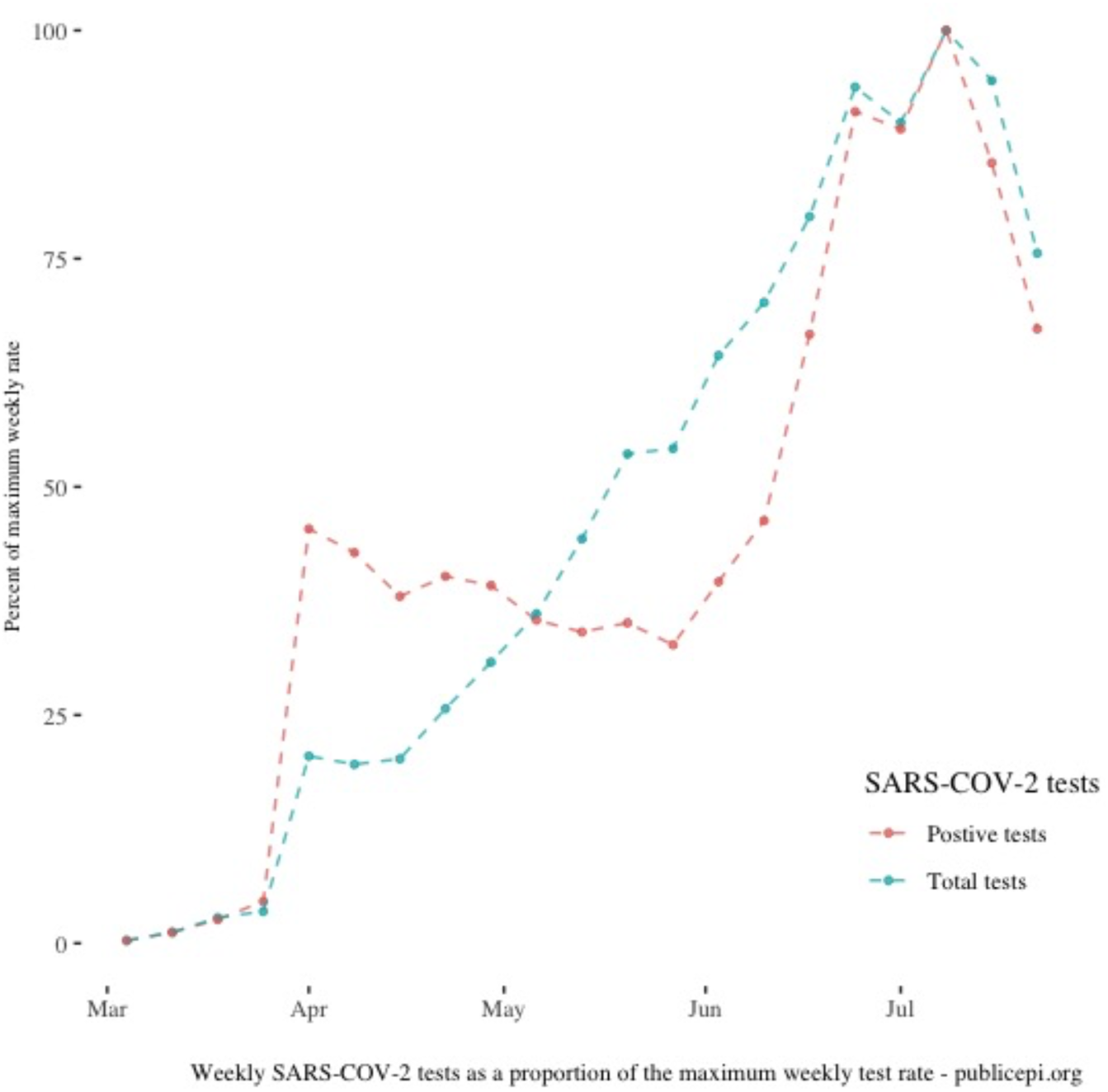
Weekly tests as a proportion of the maximum weekly tests over the SARS-COV-2 epidemic period. In April, the number of positive tests and the overall positivity rates increased sharply after FDA authorization SARS-COV-2 testing by commercial laboratories.

In the middle of a pandemic emergency, particularly one which has led to restrictions on mobility and commerce, having an unbiased and reliable measure of infectious disease incidence is not an academic issue. Optimal timing of containment and mitigation actions during a pandemic requires rigorous, consistent, and timely measures of epidemic growth available at a regional level. The 2014 update of the US CDC’s pandemic response framework identifies six interval epidemic phases: initiation, acceleration, and deceleration of epidemic transmission.^34^ Increasing epidemic growth rates define the acceleration interval of a pandemic wave while decreasing growth rates defined deceleration. Under the CDC response framework, responsive activities at Federal and State actors should be linked to epidemic intervals. Changes in case definitions and testing priorities over the epidemic period, including a nonclinical case definition, widespread asymptomatic screening, frequent and unpredictable reporting delays all make the laboratory-based SARS-COV-2 case risky for monitoring epidemic dynamics.

Hospital admission incidence rates might provide a more reliable and less biased alternative to case rates for assessment of epidemic dynamics but only if they include an objective clinical case definition. It is worth noting that in 2009, in the context of the H1N1 pandemic, CDC asked states to stop reporting of individual influenza H1N1 cases in July 2009, in part, because of the lack of clinical value of symptomatic testing. Instead of reporting cases, the CDC at the time asked States to report aggregate weekly counts of laboratory-confirmed hospitalizations or alternatively report hospitalizations meeting a syndromic definition.^1^

The observed declines in epidemic growth rates beginning in early March 2020 call into question any associations of government social distancing orders with changes in epidemic growth. These government orders occurred in rapid succession without incidence-based criteria for application and in the absence of reliable incidence data. Notably, the orders went well beyond the scope of mitigations anticipated in the US CDC’s 2017 pandemic community mitigation guidelines.^35^ State and local government actions, including those on gatherings, school closures and restrictions on commerce and mobility, largely followed secular trends including volunteer telework arrangements, abandonment of schools by concerned parents, cancellations of events and travel, and rapid population adoption of distancing behaviors. In the U.S., consumer expenditures on travel and entertainment declined beginning the week ending Feb 20^th^.^36^ By mid-March, half of Americans were avoiding public places and three-quarters avoiding public transport.^37^ Overall mobility declined by one to two-thirds depending on location.^38^

Ten regions observed by COVID-NET implemented either “stay-at-home” orders or closures of nonessential businesses in the 7-day period between March 17 and March 23, 2020. Among these regions there was significant heterogeneity both in the observed initial growth rate as well as the timing of the maximum hospitalization incidence rate in the first wave. Neither measures of growth based on hospital admissions or mortality appears influenced by the orders or their relative timing.

In summary, declining epidemic growth rates of SARS-COV-2 infection appeared in early March in the first observations of nationwide hospital admissions surveillance program in multiple U.S. regions. This finding suggests that the initiation and acceleration phases of the SARS-COV-2 pandemic occurred before February 18, 2020 at a time when US case surveillance criteria did not identify community transmission. Considering these observations along with evidence of asymptomatic carriage, presymptomatic and asymptomatic transmission, and non-specific clinical presentation, and a low clinical fraction in the young, a sizable fraction of the U.S. population may have been infected with SARS-COV-2 without detection and therefore may no longer be susceptible.

Given the societal impact of educational, economic and mobility restrictions, the possibility of a large and unobserved silent fraction of the first wave of the COVID-19 pandemic deserves further scrutiny. Timed seroprevalence studies that reflect current understanding of the antibody response and forensic epidemiology of hospitalization and death records are two ways examine this hypothesis and gain a more accurate picture of early epidemic dynamics. To more accurately monitor epidemic trends and inform pandemic mitigation planning going forward, the US CDC needs measures of epidemic disease incidence that better reflect clinical disease and account for large variations in case ascertainment strategies over time.

## Data Availability

All analysis used publicly available data

https://gis.cdc.gov/grasp/covidnet/COVID19_3.html

https://github.com/nytimes/covid-19-data

https://www.cdc.gov/coronavirus/2019-ncov/covid-data/covidview/index.html

## Acknowledgements

Appreciation for thoughtful advice of Jeffrey Klausner, Manasi Rana Suneeta Krishnan, Allan Smith and other anonymous reviewers.

